# Humanitarian Perspectives on Civilian Shelter Guidance in Armed Conflict Zones: A Qualitative Study

**DOI:** 10.1101/2025.11.06.25339465

**Authors:** Ken Brackstone, Jack Denny, Alexandra C. Hansen, Micah L. Trautwein, Elsara Badri, Aparna Cheran, Robin Toal, Serhii Tertyshnyi, Akeza A. Asgedom, Ahmed Mokhallalati, Hannah B. Wild

## Abstract

**Objectives:** The widespread use of explosive weapons in populated areas (EWIPA) has become a defining feature of modern conflict, with devastating consequences for civilians. Practical guidance on civilian sheltering during explosive attacks remains limited, inconsistent, and lacking in scientific evidence. This study explored the current landscape of shelter guidance through the perspectives of humanitarian practitioners working in EWIPA contexts.

**Methods:** Semi-structured interviews were conducted with 10 practitioners from international NGOs, Red Cross societies, and agencies engaged in risk education, emergency response, and conflict monitoring. Participants were purposively selected for operational experience in EWIPA-affected regions. Interviews explored four domains: guidance content, information sources, dissemination channels, and implementation challenges. Data were analyzed using a hybrid inductive-deductive approach.

**Results:** Practitioners described a range of sheltering messages, from general cues like “find cover” to specific techniques including low-profile positioning. Most guidance drew on field experience and expert consensus rather than empirical research. Dissemination strategies varied by context. Challenges included message distortion, difficulty engaging high-risk groups, and the absence of standardized recommendations.

**Conclusions:** Current civilian shelter guidance in EWIPA contexts is fragmented and lacks an evidence base. Findings highlight the need for coordinated, context-specific, and evidence-informed approaches to strengthen civilian protection.

## Introduction

Armed conflict has undergone a dramatic transformation over the last decade. Once limited to the battlefield with clearly defined front lines, modern warfare has become increasingly urbanised, asymmetric, and protracted. Armed conflict is now characterised by blurred combat zones, the involvement of non-state actors, and the frequent targeting of civilian infrastructure [1]. This transformation is marked not only by the proliferation of conflict, but also due to the military’s increased use of high precision and high-energy weapons. The increasing use of explosive weapons in populated areas (EWIPA) has emerged as a defining feature of modern warfare, with widespread and devastating consequences for civilians [2]. Action on Armed Violence (AOAV) documented 47,476 causalities from explosive weapons in 2023 alone, with civilians accounting for 90% of casualties recorded in populated areas [3]. Civilian deaths increased by 130% compared with 2022, largely due to conflicts in Gaza, Sudan, and Myanmar [3]. Complementary monitoring data recorded 8,826 incidents of explosive violence between August 2023 and July 2024, resulting in 52,050 civilian casualties in regions including Palestine, Myanmar, Pakistan, Sudan, Syria, and Ukraine [4–6]. Civilians accounted for 85% of total casualties during this period, highlighting both the global scale of EWIPA-related harm and disproportionate burden borne by civilians. Consequences are often profound with attacks resulting in simultaneous injuries or deaths of large numbers of people within a single strike. Communities are displaced, health and education systems are destroyed, and psychological trauma becomes widespread and long-lasting [7]. There is therefore an urgent need for protective measures tailored toward civilians, and for new frameworks that ensure safe locations and protection of populations caught in the crossfire.

The International Committee of the Red Cross (ICRC) has warned that the indiscriminate effects of EWIPA violates principles of international humanitarian law [8]. In response, the Political Declaration on Strengthening the Protection of Civilians from the Humanitarian Consequences of Explosive Weapons in Populated Areas – endorsed by over 80 countries as of 2023 – has aimed to raise awareness of EWIPA’s humanitarian effects and establish new international standards for protecting civilians from explosive weapons in populated areas [9]. The Sendai Framework for Disaster Risk Reduction similarly emphasizes the importance of both structural and non-structural measures in protecting at-risk populations [10]. Non-structural interventions such as risk communication and protective messaging has become essential components of civilian protection in conflict settings where formal shelters may be inaccessible. Despite their relevance, the development and delivery of such guidance remain under-examined and are rarely grounded in empirical evidence, and casualty numbers from the use of EWIPA continue to rise [3].

In many active conflict zones, civilian populations often lack access to practical guidance on how to shelter during aerial bombardments, shelling, and other explosive attacks [11]. A scoping review recently conducted by the Explosive Weapons Trauma Care Collective (EXTRACCT) found a significant lack of standardized or empirically validated shelter recommendations designed for civilians exposed to EWIPA [11]. Few studies have rigorously evaluated which sheltering strategies are most effective under different explosive scenarios. Many were originally developed for military personnel or post-conflict scenarios, and may not translate to the realities faced by civilians in urbanised warzones. There is also little standardized, evidence-based guidance readily available for civilians on the ground [11]. Often, civilians rely on word of mouth, intuition, or unverified information circulating on social media. For example, civilians in Gaza turned to platforms such as X (formerly Twitter) to share survival strategies during shelling. Individuals previously posted recommendations about which floors of a building offered better protection, how to reinforce shelter using household materials, and where to position during an airstrike. However, while these practices demonstrate resilience and community solidarity, they reveal gaps in institutional support. In many low-income countries affected by recurring conflicts, access to even informal survival techniques or digital information-sharing may be limited, leaving civilians with no reliable sources of protection guidance. The accuracy and effectiveness of this crowd-sourced advice are unknown, and in some cases, may be misleading or harmful [11].

Further research is needed to evaluate current shelter guidance available to civilians and determine the extent to which these recommendations are evidence-based. It is equally important to investigate how such guidance is disseminated by humanitarian actors across organizations and regions. Our primary aim was to conduct a qualitative assessment of shelter guidance currently provided to civilians during explosive violence from the perspective of humanitarian experts and practitioners working in conflict-affected settings. We conducted interviews with individuals from key humanitarian sectors including emergency response, civilian protection, public health, and mine action groups to document sheltering recommendations currently being given, identify gaps or inconsistencies in existing guidance, and assess whether these recommendations are based on empirical evidence or field-based experience. These findings may inform future efforts to develop more standardized, accessible, and evidence-based shelter guidance for populations affected by EWIPA.

## Methods

### Recruitment and data collection

We conducted once-off in-depth interviews of 30-60 minutes with 10 key stakeholders of humanitarian organizations in relevant domains such as risk reduction, injury prevention, explosive ordnance risk education, conflict preparedness, and protection. Participants were emailed and interviews were conducted between February and April 2024.

We applied maximum variation purposive sampling to include interviewees working in humanitarian or protection-focused organizations who had experience working in combat settings affected by EWIPA for the last 5-10 years. Eligible participants were senior or technical officers with experience in a wide range of settings. Specifically, we sought stakeholders from international and regional humanitarian organizations, including NGOs and UN agencies. Organizations included the Norwegian People’s Aid (NPA), Humanity and Inclusion (HI), Mines Advisory Group (MAG), Geneva International Centre for Humanitarian Demining (GICHD), United Nations Development Programme (UNDP), and ICRC. We sought participants with direct experience advising on, implementing, or disseminating sheltering guidance for civilians in conflict-affected areas due to military actions.

Participants were sought through established contacts and international collaborators. We detailed the nature of the study to potential participants. We invited 14 stakeholders to participate, but were unable to arrange interviews with 4 of these due to availability constraints. Semi-structured interviews were conducted online using Zoom, with all sessions hosted over a secure VPN. Interviews were conducted by HW and EB, both female researchers (a clinician and researcher, respectively), with training in qualitative methods and experience in emergency care and humanitarian response in conflict settings. Interviews were audio-recorded. Only individual participants and both researchers were present in interviews, and no field notes were made. Verbal informed consent was obtained prior to each interview.

### Data collection tools

Participants first completed a personal information form, collecting details about their current organization, and professional role and area(s) of expertise (Table 1). Other information was gathered but has been omitted from reporting to protect participants’ identities, especially given the small sample size and close-knit nature of humanitarian mine action and protection sectors. A semi-structured interview guide was developed based on a literature review and pilot-tested with a second researcher to ensure clarity and appropriateness (Table 2). The interview guide consisted of seven key open-ended questions, and was focused on four domains: (1) nature and content of shelter guidance provided to civilians; (2) sources and development processes of such advice; (3) dissemination channels and formats used to communicate guidance, and (4) barriers and challenges to effective civilian sheltering during explosive violence. The interview codebook is included in Appendices.

**Table 1.** Summary of interviewed sector experts.

| Participant no. | Organisation | Role |
| --- | --- | --- |
| 1 | International NGO | Senior advisor in humanitarian operations |
| 2 | International research and training centre | Programme management specialist |
| 3 | International NGO | Technical specialist in humanitarian programming |
| 4 | International NGO | Advisor in conflict response |
| 5 | International NGO | Specialist in programme monitoring |
| 6 | Humanitarian organisation | Humanitarian operations specialist |
| 7 | Humanitarian organisation | Technical expert in weapons contamination |
| 8 | International NGO | Field operations and community engagement specialist |
| 9 | United Nations Agency | Advisor in humanitarian mine action |
| 10 | International NGO | Policy and advocacy specialist |

**Table 2.** The semi-structured interview guide.

| Questions |
| --- |
| 1. Can you describe the key types of advice or guidance your organization provides to civilians sheltering during explosive attacks in populated areas? |
| 2. How does your organization usually decide what guidance to give? |
| 3. Could you walk me through how this shelter guidance typically reaches civilians in the field? |
| 4. What forms of media or materials are most commonly used to communicate these messages to affected populations? |
| 5. In your experience, what changes or improvements are needed to strengthen the quality or impact of current sheltering guidance for civilians in EWIPA settings? |
| 6. What kinds of challenges have you observed when trying to deliver or implement guidance in real-world conflict settings due to military actions? |

### Data preparation and analysis

Interviews were transcribed verbatim, de-identified, and checked for completeness and accuracy. Transcripts were stored on an encrypted, password-protected server accessible only to the research team. Analysis was conducted using a combination of deductive and inductive approaches. Deductive codes were drawn from the interview guide and research objectives, while inductive codes were derived from emerging data patterns [12,13]. The interview guide was designed to explore key operational dimensions of civilian shelter guidance in EWIPA settings. As such, domains of shelter guidance, information sources, dissemination channels, and implementation challenges were embedded in the structure of questions. Coding was carried out using Dedoose software (version 9.0.62) [14]. Two researchers independently coded transcripts using a shared codebook, and discrepancies were resolved through iterative review and consensus-building sessions. An initial coding framework was developed and refined based on early transcripts. Participants did not provide feedback on findings due to the time-sensitive nature of data collection and challenges of recontacting participants in humanitarian and conflict settings.

### Ethical considerations

The study was reviewed and approved by the University of Washington Human Subjects Division (IRB ID: STUDY00019413). All participants provided informed verbal consent before interviews commenced. Participants were assured of confidentiality and the voluntary nature of their participation. No identifying information was included or reported in findings. Audio recordings and transcripts were securely stored and accessible only to the research team.

### Trustworthiness and rigour

To ensure credibility, the interview guide was pilot-tested, and a standardized protocol was followed across interviews. Triangulation was achieved through the use of multiple coders and iterative discussion of findings (AH, MT). Member checking was conducted by sharing emerging interpretations with selected participants to validate accuracy. Participant quotations are presented in a table to enhance transparency.

## Results

Ten experts were interviewed across six organizations working in conflict-affected settings. These individuals brought extensive operational experience from diverse conflict-affected contexts, including Gaza, Lebanon, Myanmar, Ukraine, and Yemen, and many had contributed directly to the development or dissemination of shelter guidance at global or regional levels. Analyses were organized around four key domains reflecting the structure of the interview guide and participants’ detailed responses: (1) nature and content of shelter guidance provided to civilians; (2) sources and development processes of such advice; (3) dissemination channels and formats used to communicate guidance, and (4) barriers and challenges to effective civilian sheltering during explosive violence. See Table 3 for representative quotes by domains of enquiry.

**Table 3.**
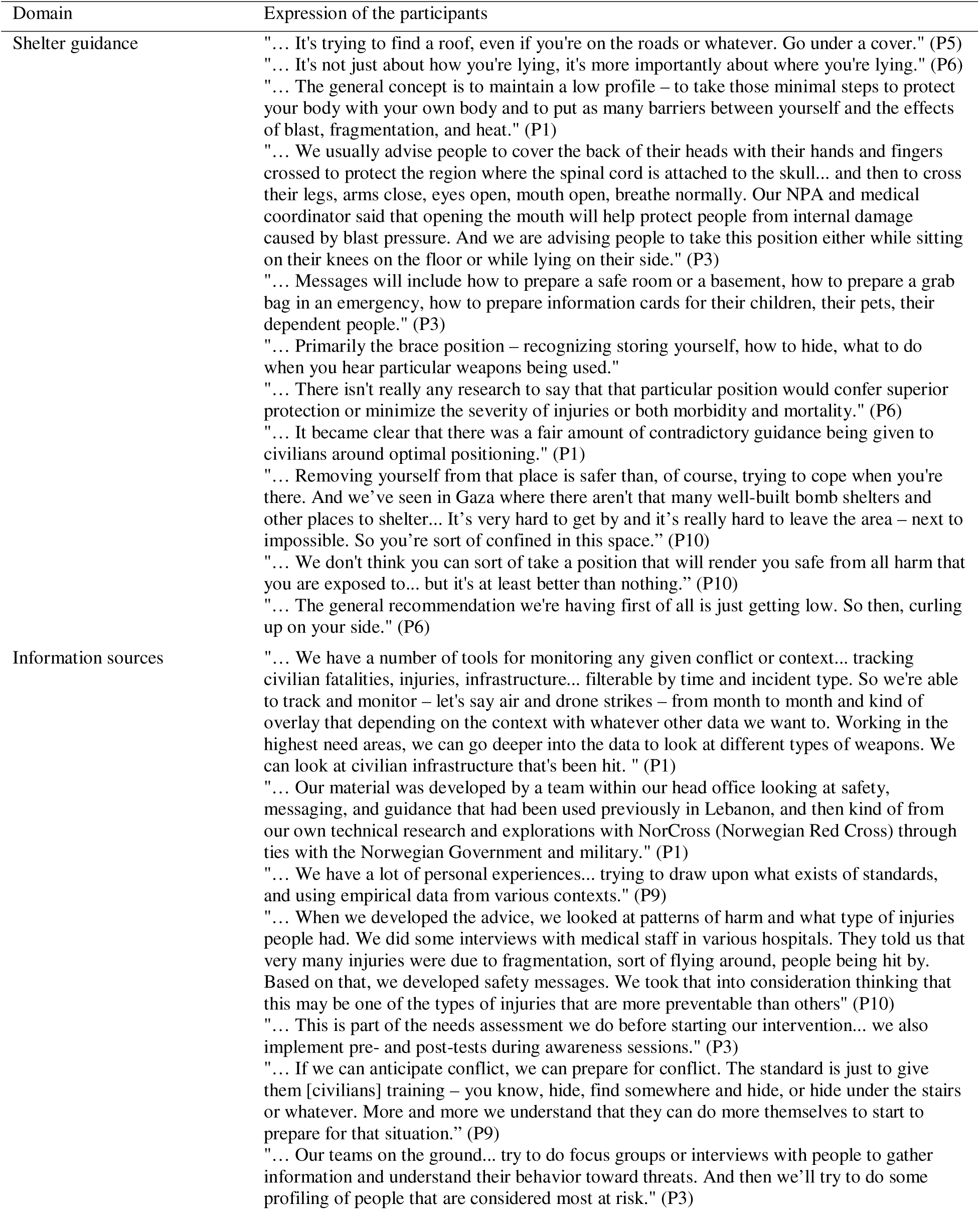
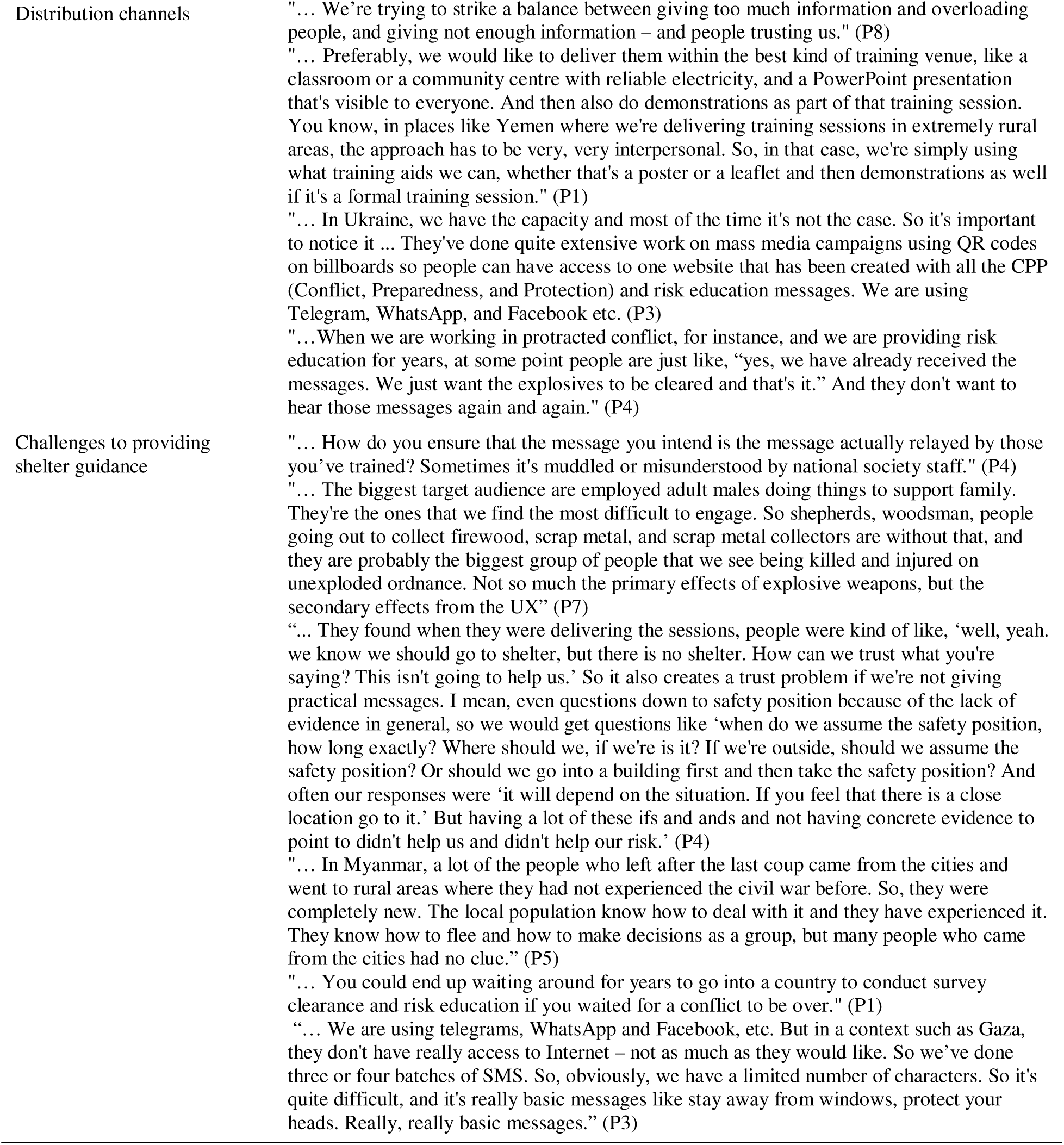
Expression of participants.

### Domain 1. Shelter guidance

Experts consistently emphasised the importance of conveying practical and actionable information about protective behaviors during explosive attacks. Key recommendations included seeking physical cover, maintaining a low profile, and assuming specific body positions to reduce risks of injury from blast, fragmentation, and heat. However, there was considerable variation in the level of detail provided across different contexts, with some agencies offering more general advice (e.g., “get low”, or “find cover”), with others recommending more specific postures and protective techniques. Notably, several experts acknowledged a lack of empirical evidence supporting any one optimal position, leading to consistencies in guidance across agencies and settings. Shelter guidance offered was described as “better than nothing” in high-risk areas where civilians are unable to evacuate – not a guarantee of safety, but a harm-reduction strategy.

### Domain 2. Information sources

The content of shelter guidance was informed by a combination of field observations, retrospective injury data, expert judgment, and operational experience. Some organizations reported incorporating empirical insights from conflict-specific injury patterns (e.g., the prevalence of fragmentation injuries) into the development of their messages. In other cases, pre-existing protocols from previous conflicts were adapted to new contexts. Experts also described drawing on needs assessments, including community interviews and pre/post evaluations of safety training, to tailor messaging to specific risks faced by civilian populations. The iterative nature of guidance development was a common theme across interviews, such as combining existing standards, local feedback, and contextualized data.

### Domain 3. Distribution channels

Dissemination of shelter guidance varied widely depending on access, infrastructure, and conflict stage. In optimal scenarios, agencies aimed to deliver structured training in community centers or classrooms with visual materials and in-person demonstrations. However, distribution relied on informal methods such as posters, printed leaflets, and interpersonal communication in active or rural conflict zones. Digital platforms were also used in contexts like Ukraine, where technological infrastructure permitted mass media campaigns (e.g., messaging apps, websites, and QR codes on public signage). Despite these efforts, message fatigue was reported in prolonged crises. where civilians expressed frustration at repeated exposure to safety messaging without corresponding progress on demining or protection measures.

### Domain 4. Challenges to providing shelter guidance

Experts identified practical and contextual challenges in delivering effective shelter guidance. A recurrent concern related to the potential for messages to be misunderstood or diluted when relayed by local trainers or partner organizations, especially in settings with limited oversight or high staff turnover. Difficulties also arose in engaging high-risk subpopulations, especially adult males engaged in outdoor labor who were perceived as the least receptive to training but among the most vulnerable to injury. Furthermore, displaced populations lacked generational or community knowledge that might otherwise aid in self-protection, such as urban residents who had newly arrived in rural conflict zones. Finally, several experts acknowledged logistical barriers to delivering timely guidance during ongoing conflict, including security risks, funding constraints, and delays in obtaining access to affected regions.

## Discussion

This qualitative study aimed to clarify how humanitarian actors conceptualize protective sheltering and inform future efforts toward more standardized, accessible, and evidence-based guidance for populations at risk from EWIPA. Through interviews with expert practitioners engaged in risk education, civilian protection, and injury surveillance, four domains were assessed: (1) nature and content of shelter guidance; (2) its sources and development; (3) dissemination channels and formats; and (4) barriers to effective civilian sheltering during explosive violence.

Shelter messaging was a core protection component across settings. Participants noted that simple actions – finding cover, staying low, and shielding vulnerable body parts – can reduce harm, though they do not guarantee safety. As one participant wrote: “We don’t think you can [sort of] take a position that will render you safe from all harm… but it’s at least better than nothing” (P10). Such strategies are crucial where evacuations or shelters are unavailable, particularly regions most affected by explosive violence such as densely populated or besieged areas like Gaza or eastern Ukraine [5,7]. Guidance varied in specificity: some organisations advised only “get low” or “find cover,” while others suggested bracing conditions or protecting the next and head. These techniques were based on consensus rather than research, and participants acknowledged that lack of evidence. This variability led to inconsistent and contradictory advice. EXTRACCT’s scoping review similarly identified a lack of standardized, evidence-informed guidance tailored for civilians [11]. Coordinated research into biomechanics and optimal posters are needed, as most existing studies focus on military contexts [11].

The absence of harmonized guidance may undermine both effectiveness and public trust. Participants described instances in which conflicting recommendations, such as variations in body positioning or safe-room preparation, created confusion among affected populations. In conflict settings where institutional trust is often fragile and misinformation is widespread, such inconsistencies may be perceived as indicative of uncertainty or unreliability. This, in turn, may reduce uptake of otherwise life-preserving guidance or lead individuals to prioritise informal or unverified sources of information. A contemporary example that illustrates these dynamics was the conflict in Gaza. Despite the scale of bombardment, civilians often lacked access to designated shelters or fortified buildings, forcing families to remain in homes that provide little protection against modern munitions. Over 60% of buildings were destroyed in certain parts of the region, displacing a vast number of people into tents or overcrowded schools that offered almost no protection [15]. United Nations Office for the Coordination of Humanitarian Affairs (OCHA) further estimates that nearly 1.9m people – about 85% of Gaza’s population – were displaced, with a large proportion living in temporary shelters lacking basic safety standards [16]. No formal alarm or warning system existed to shelter, and when warnings were issued by Israeli forces, they often contained errors and were ineffective due to widespread destruction and overcrowding [17]. Thus, bombardments occurred continuously and indiscriminately, and offer no time for protective responses.

The development of shelter guidance drew from a range of sources, including field observations, retrospective injury surveillance, expert judgement, and needs assessments with affected communities. No participants reported having empirical evidence from which to develop shelter guidance. In some settings, conflict-specific data informed recommendations. For example, training materials were adapted accordingly to emphasise protective behaviors against shrapnel when reported that fragmentation injuries were the most common blast-related trauma in a given area. However, the process of evidence integration was not systematic across all organizations. Some participants described drawing on protocols from past conflicts and adapting them for use in new emergencies, whereas others relied on staff with operational memory and technical experience. These variations emphasise concerns raised by humanitarian organizations regarding the translation of military or post-conflict guidelines into current conflict zones [10]. Some organizations have adopted more structured approaches and conducted pre-intervention assessments and post-training evaluations to refine their messages, but others utilise focus groups to tailor advice to specific population groups or risk behaviors. These findings highlight the importance of integrating local knowledge, conflict-specific data, and systematic assessment methods into message development, especially given that over 85% of civilian casualties from EWIPA in 2023 occurred in just a handful of conflict settings [5].

The dissemination of shelter guidance varied depending on the security situation, infrastructure, and conflict stage. In well-resourced contexts like Ukraine, agencies delivered multimedia risk education via websites, messaging apps, and QR codes displayed in public spaces. In contrast, in low-resource or high-risk settings, guidance was often distributed through interpersonal methods, including community trainers, posters, or oral demonstrations. This diversity reflects not only contextual constraints, but strategic trade-offs. Digital platforms can reach large audiences rapidly, but require stable internet access and trust in the information source [18]. While community-generated content demonstrates civilian solidarity, it highlights a lack of trusted, centralized sources of information. By contrast, interpersonal delivery allows for clarification and adaptation to audience needs, but is slower and more resource-intensive. Message fatigue also emerged as a concern in protracted emergencies, especially where repeated exposure to protection messages without visible impact led to frustration or disengagement [19]. These findings align with established principles of risk communication, which highlight clarity, credibility, and contextual relevance as essential for effective protective messaging [19].

Challenges included message distortion during secondary delivery, particularly when guidance was relayed by local trainers with limited supervision. Risks of dilution or misinterpretation are heightened in contexts of rapid staff turnover, limited training, or political sensitivities. Reaching high-risk groups also proved difficult. Adult men engaged in outdoor or manual labor were seen as especially vulnerable to blast injury, but often least engaged with training. Structural factors such as gender roles, occupational norms, and mistrust of outsiders contributed to this disconnect [20]. Displaced populations unfamiliar with local protective practices faced similar challenges, as seen in Myanmar, where urban residents relocated to rural areas without generational knowledge of how to respond to armed violence. These issues echo wider difficulties noted by the ICRC and UN, where mobility, displacement, and fragmented authority can limit preparedness [7,9]. Operational barriers further complicate delivery, including security risks, bureaucratic restrictions, and funding constraints. In some cases, organizations waited months or years for permission to conduct risk education. Participants also questioned the credibility of advice such as “go to shelter” in contexts with no safe shelters, while educators felt underprepared to answer detailed questions due to limited evidence. Communication constraints added barriers, particularly in Gaza, where poor internet access forced reliance on SMS campaigns that reduced messages to the most basic advice. These issues illustrate how both the form and substance of safety guidance can undermine its effectiveness.

### Implications for policy and practice

There is a strong need for greater harmonization of protective advice across organizations and contexts, informed by emerging evidence on injury mechanisms and environmental factors. Inconsistent guidance can lead to public confusion, reduce trust, and undermine uptake [21]. Coordinated messaging that is grounded in biomechanical and clinical understanding, conflict-specific injury patterns, and local feedback, may help to address these challenges. The development of a central, regularly updated library of sheltering recommendations may offer a useful starting point for harmonization efforts. These may be tailored to specific weapons, settings, and population groups. Existing bodies within humanitarian mine action groups have the potential to play an important role in such coordination.

While international organizations such as the ICRC and United Nations High Commissioner for Refugees (UNHCR) have developed guidelines for sheltering in urban warfare contexts, these recommendations often lack practical detail needed for effective on-the-ground application. For instance, the ICRC’s *Reducing Civilian Harm in Urban Warfare: A Commander’s Handbook* provides strategic guidance for military commanders, but offers limited actionable advice for civilians seeking shelter during active conflict zones [22]. Similarly, UNHCR’s *Emergency Shelter Solutions and Standards* outlines minimum space requirements and structural considerations, but does not delve into specific sheltering strategies in urban warfare zones [23]. In this regard, while Political Declaration of EWIPA outlines a list of pledges that aim to mitigate civilian harm by strengthening national policies, supporting victims, and improving data collection on the effects of EWIPA, such diplomacy and policy must be paired with practical shelter guidelines to reduce civilian harm. Given these gaps, our findings reinforce the need for organizations to advocate for the prevention and lawful restriction of EWIPA, and to enhance their guidance by incorporating detailed, context-specific sheltering strategies that address the immediate needs of civilians on conflict settings.

Communication strategies must be carefully designed to reflect the information ecosystem of each conflict setting. Tools such as mobile apps, QR codes, and social media can enhance reach and speed of dissemination in digitally connected environments. However, in low-resource or remote areas, interpersonal communication via trusted community actors remains vital [20]. Hybrid dissemination strategies such as combining mass communication with in-person engagement may be effective in reducing messaging saturation and ensuring comprehension. Incorporating mechanisms for monitoring, evaluation, accountability, and learning (MEAL), including pre- and post-assessments and structured community feedback, is also critical to ensure that guidance remains effective and responsible to local needs.

Further, there is a clear need for capacity building. Training programmes should equip frontline staff, including local volunteers and community responders, with up-to-date knowledge and communication skills. These individuals are often the first point of contact for at-risk populations, and their ability to deliver accurate, context-appropriate shelter advice is critical. Training should also emphasize the importance of identifying information gaps and reporting them to relevant agencies for iterative improvement. Finally, shelter guidance should be embedded within broader civilian protection strategies that includes explosive ordnance risk education, infrastructure support, psychosocial assistance, and advocacy for adherence to international humanitarian law. In many conflict-affected settings, shelter guidance may be one of the few protective actions available to civilians during acute phases of violence. Its effectiveness will depend not only on content, but also on delivery, trust, and integration with complementary forms of support.

### Strengths and limitations

A key strength of this study is its inclusion of practitioners with a diverse range of expertise and operational experience across multiple conflict-affected regions. The semi-structured format also allowed for nuanced accounts that illuminated the reasoning behind strategic decisions in the field. However, the sample was relatively small and relied partly on snowball sampling. Second, the study focused exclusively on the perspectives of staff from international humanitarian NGOs and did not include insights from local organisations or civilians themselves. As such, the findings may not capture context-specific knowledge of local actors, or the lived experiences of civilians who are the intended users of shelter guidance. This is particularly relevant in regions such as parts of Africa, where recurrent conflict and resource limitations may foster unique sheltering practices and survival strategies that differ from those observed in other settings. Future research that incorporates local organisations and civilian perspectives could provide regionally grounded insights and more contextually tailored recommendations for shelter guidance.

## Conclusion

Civilians living in conflict zones face acute risk from explosive weapons, yet evidence-based shelter guidance remains largely limited. While such guidance cannot eliminate harm, it can provide a meaningful injury prevention strategy. More research is urgently needed to strengthen recommendations, while improved standardization is essential. Fragmented guidance risks undermine trust and uptake. Messages must adapt to structural conditions, weapon types, and the needs of vulnerable groups through long-term partnerships. Alongside policy-level efforts to restrict EWIPA, accurate and standardized shelter guidance is a humanitarian obligation.

## Supporting information

COREQ checklist

## Data Availability

The data are not publicly available because participants did not consent to the sharing of their interview transcripts or recordings beyond the research team.

## Author contribution

KB: Writing of first draft, manuscript revisions; JD: Writing of first draft, manuscript revisions; ACH: Protocol development, data analysis, and manuscript revisions; MLT: Protocol development, data analysis, and manuscript revisions; EB: Protocol development, data collection, data analysis, and writing of first draft; AC: Protocol development and data analysis; RT: data analysis and writing of first draft; ST: manuscript revisions; AAA: manuscript revisions; AM: manuscript revisions; HW: Conception, supervision, data analysis, data extraction, manuscript revisions.

## Competing interests

No competing interests declared.

## Appendix

Interview Codebook

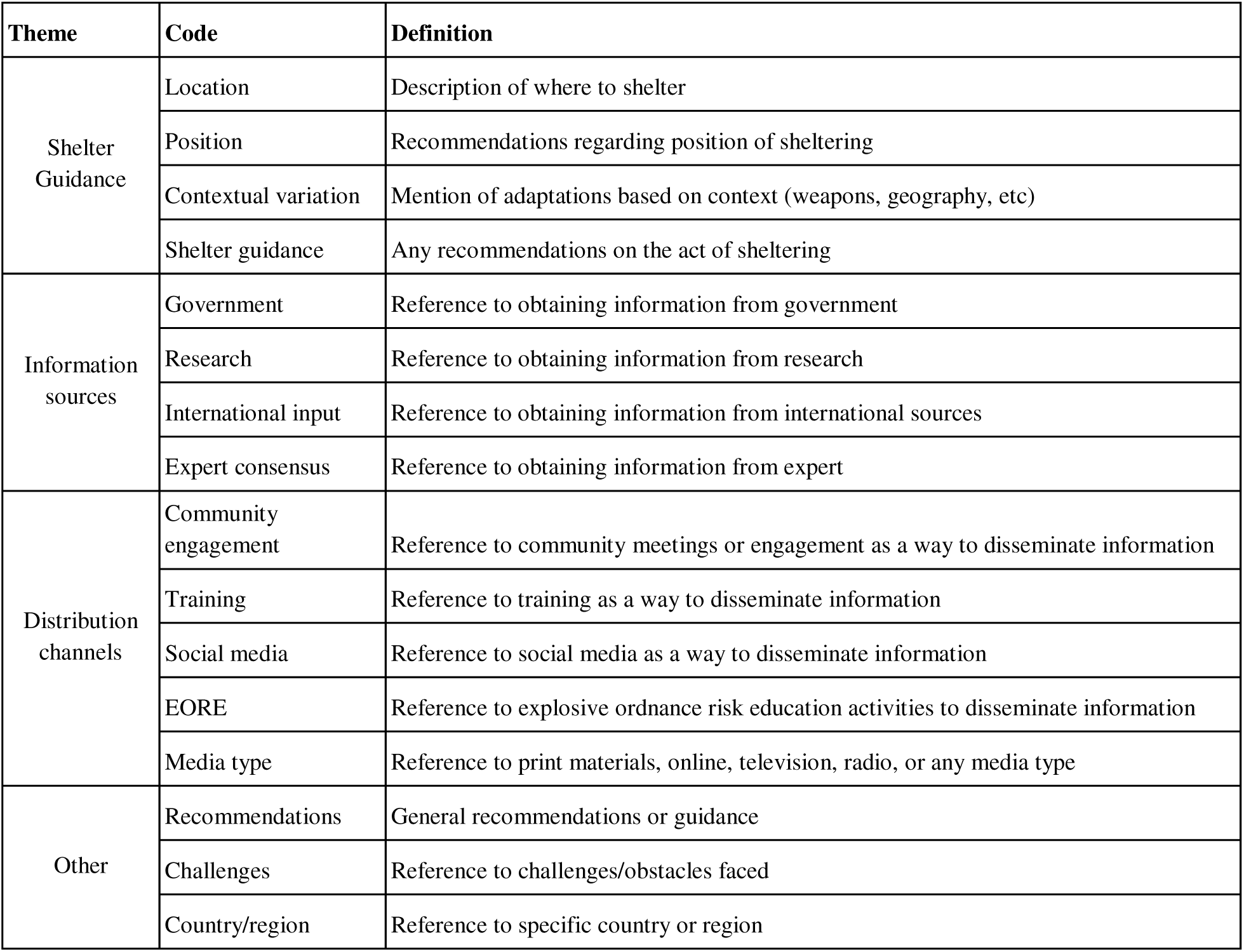

| Theme | Code | Definition |
| --- | --- | --- |
| Shelter Guidance | Location | Description of where to shelter |
|  | Position | Recommendations regarding position of sheltering |
|  | Contextual variation | Mention of adaptations based on context (weapons, geography, etc) |
|  | Shelter guidance | Any recommendations on the act of sheltering |
| Information sources | Government | Reference to obtaining information from government |
|  | Research | Reference to obtaining information from research |
|  | International input | Reference to obtaining information from international sources |
|  | Expert consensus | Reference to obtaining information from expert |
| Distribution channels | Community engagement | Reference to community meetings or engagement as a way to disseminate information |
|  | Training | Reference to training as a way to disseminate information |
|  | Social media | Reference to social media as a way to disseminate information |
|  | EORE | Reference to explosive ordnance risk education activities to disseminate information |
|  | Media type | Reference to print materials, online, television, radio, or any media type |
| Other | Recommendations | General recommendations or guidance |
|  | Challenges | Reference to challenges/obstacles faced |
|  | Country/region | Reference to specific country or region |

## Notes

### Competing Interest Statement

The authors have declared no competing interest.

### Funding Statement

This study did not receive any funding.

### Author Declarations

The study was reviewed and approved by the University of Washington Human Subjects Division (IRB ID: STUDY00019413).

## References

1. United Nations. With highest number of violent conflicts since Second World War, United Nations must rethink efforts to achieve, sustain peace, speakers tell Security Council. Meetings Coverage and Press Releases. Published 26 January, 2023. Accessed 19 September, 2025. https://press.un.org/en/2023/sc15184.doc.htm

2. Policinski E, Kuzmanovic J. Protracted conflicts: The enduring legacy of endless war. Int Rev Red Cross. Published November, 2019. Accessed 16 September, 2025. http://international-review.icrc.org/articles/protracted-conflicts-enduring-legacy-endless-war-ir912

3. AOAV. Explosive Violence Monitor: AOAV. Published April, 2024. Accessed September 19, 2025. https://aoav.org.uk/2024/explosive-violence-monitor-2023

4. International Committee of the Red Cross. ICRC president: “We are witnessing a global and collective failure to protect civilians in armed conflicts”. Published 22 April, 2024. Accessed 15 September, 2025. 2https://www.icrc.org/en/document/global-and-collective-failure-to-protect-civilians-in-armed-conflict

5. Explosive Weapons Monitor. Explosive Weapons Monitor. Accessed 29 August, 2025. https://explosiveweaponsmonitor.org

6. International Network on Explosive Weapons. Protecting civilians from the use of explosive weapons in populated areas. Published October, 2024. Accessed 29 August, 2025. https://www.inew.org/wp-content/uploads/2024/10/inew_oct_protecting_24_v2.pdf

7. Brackstone K, Denny J, Wild H, et al. Landmine exposure and hearing impairment among war-affected Ukrainian civilians: Exploring the role of PTSD as a contributory pathway. Under review.

8. International Committee of the Red Cross. Explosive weapons with wide area effects: A deadly choice in populated areas. 2022. Accessed 29 August, 2025. https://www.icrc.org/sites/default/files/document_new/file_list/ewipa_explosive_weapons_with_wide_area_effect_final.pdf.

9. United Nations Office for Disarmament Affairs. The political declaration. Accessed 29 August, 2025. https://ewipa.org/the-political-declaration.

10. United Nations Office for Disaster Risk Reduction. What is the Sendai Framework for Disaster Risk Reduction? Accessed 29 August, 2025. https://www.undrr.org/implementing-sendai-framework/what-sendai-framework

11. Hansen AC, Badri E, Almalla M, et al. Civilian sheltering guidelines for explosive weapons in populated areas: A scoping review. Disaster Med Pub Health Prep. 2025;19. doi: 10.1017/dmp.2025.117

12. Fereday J, Muir-Cochrane E. Demonstrating rigor using thematic analysis: A hybrid approach of inductive and deductive coding and theme development. Int J Qual Methods. 2006;5:80–92. doi: 10.1177/160940690600500107.

13. Tong A, Sainsbury P, Craig C. Consolidated criteria for reporting qualitative research (COREQ): a 32-item checklist for interviews and focus groups, Int J for Qual in Healt Car. 2007;19(6):349–357. doi: 10.1093/intqhc/mzm042.

14. Dedoose. Dedoose makes qualitative and mixed methods analysis easy. Accessed 19 August, 2025. https://www.dedoose.com/

15. Scher C, Leventhal MP, Van Den Hoek J. Active InSAR monitoring of building damage in Gaza during the Israel-Hamas War. arXiv. doi: 10.48550/arXiv.2506.14730.

16. Human Rights Watch. “Hopeless, starving, and besieged”: Israel’s forced displacement of Palestinians in Gaza. Published November 14, 2024. Accessed 29 August, 2025. https://www.hrw.org/report/2024/11/14/hopeless-starving-and-besieged/israels-forced-displacement-palestinians-gaza

17. United Nations Office for the Coordination of Humanitarian Affairs – oPt. Humanitarian situation update #275: Gaza Strip. Published 25 March, 2025. Accessed 29 August, 2025. https://www.ochaopt.org/content/humanitarian-situation-update-275-gaza-strip

18. West DM. Digital divide: Improving Internet access in the developing world through affordable services and diverse content. Published February 2015. Accessed 29 August, 2025. https://www.brookings.edu/wp-content/uploads/2016/06/West_Internet-Access.pdf

19. Baseman JG, Revere D, Painter I, et al. Public health communications and alert fatigue. BMC Health Serv Res. 2013;13(295).

20. Cojocaru V, Dincu I. CRVS communication in emergency, conflict-affected, and fragile settings. Accessed 29 August, 2025. https://idl-bnc-idrc.dspacedirect.org/server/api/core/bitstreams/4c7aa8cb-40f8-45ff-b20b-9b3c350a90ba/content

21. Covello VT, Sandman PM. Risk communication: Evolution and revolution. In AB Wolbarst (Ed.). Solutions to an Environment in Peril. 2001:164–178. John Hopkins University Press.

22. International Committee of the Red Cross. Reducing civilian harm in urban warfare: A commander’s handbook. Published November, 2021. Accessed 29 August, 2025. https://www.icrc.org/en/document/reducing-civilian-harm-urban-warfare-commanders-handbook

23. United Nations High Commissioner for Refugees. Emergency shelter solutions and standards. Published January 30, 2025. Accessed 25 August, 2025. https://emergency.unhcr.org/emergency-assistance/shelter-camp-and-settlement/shelter-and-housing/emergency-shelter-solutions-and-standards

